# ArchSpiral: An iPad-Based Digital Archimedean Spiral Assessment for Objective Motor Evaluation in Rare Diseases

**DOI:** 10.64898/2026.07.27.26358872

**Authors:** Daniel R. Schecter, Rory J. Tinker, Matteo Danieletto, Georgia MacDonald, Tamas Kozicz, Eva Morava, Benjamin S Glicksberg

**Author notes:** **CORRESPONDING AUTHOR:** Daniel Schecter. Co-senior authorship (Eva Morava, Benjamin S Glicksberg).

## Abstract

**Objectives:** We developed and evaluated ArchSpiral, an iPad application that digitizes the International Cooperative Ataxia Rating Scale (ICARS) spiral tracing task to address the need for portable rare disease assessment while preserving clinical scoring and generating quantitative digital biomarkers.

**Materials and Methods:** ArchSpiral (Swift/SwiftUI/PencilKit) captures Apple Pencil or finger tracings and computes tracing accuracy, root mean squared error, duration, average speed, pen lifts, pauses, steadiness, and an automated ICARS-compatible morphology score. Twelve participants with congenital disorders of glycosylation completed paired paper-digital assessments.

**Results:** Digital ICARS scores spanned the scoring range (1–4). Paper and digital scores agreed exactly in 11 of 12 assessments (linear weighted κ = 0.91). Steadiness showed the strongest association with paper-based ICARS scores (ρ = −0.82, FDR-adjusted *P* = 0.008).

**Discussion:** ArchSpiral enables standardized digital spiral assessment, while preserving compatibility with conventional ICARS scoring and adding objective quantitative measures for longitudinal rare disease research and clinical care.

## Background and Significance

The Archimedean spiral, a curve whose successive turns are separated by a constant distance, is a long-standing substrate for assessing upper-limb motor function. Because loop spacing is uniform, deviations produced by tremor, dysmetria, or decomposition are apparent, making the spiral a sensitive test of fine motor control. Performed either as a freehand drawing or by tracing a pre-printed template, spiral assessment has been applied across essential and functional tremor, Parkinson disease, cerebellar ataxia, multiple sclerosis, and age-related fine motor decline.[1–6] It is also embedded in standardized clinical rating instruments, including the Bain & Findley Spirography Scale, the Fahn-Tolosa-Marin Tremor Rating Scale, and the International Cooperative Ataxia Rating Scale (ICARS).[7–9] Among these, ICARS has been used to quantify cerebellar dysfunction in rare genetic disorders, where standardized longitudinal motor assessments are essential for defining natural history, monitoring disease progression, and evaluating treatment response.[10–13]

ICARS is a validated 100-point scale for cerebellar syndrome used in the natural history protocol for congenital disorders of glycosylation (CDG), a heterogeneous group of more than 200 genetic disorders of protein and lipid glycosylation that result in complex neurologic disease characterized by ataxia and extrapyramidal symptoms, to monitor disease progression and therapeutic response. [7, 14, 15] Its kinetic function domain includes an Archimedean spiral task scored 0 (normal) to 4 (disorganized or impossible). Unlike most digital spiral applications, the ICARS task requires tracing a pre-drawn three-turn template rather than drawing freehand, a distinction central to faithful digital implementation.

Digital spiral analysis is not new: prior work has captured spiral drawings on tablets and digitizers to derive kinematic biomarkers in Parkinson disease, spinocerebellar degeneration, and multiple sclerosis.[3–6] These tools rely on freehand drawing and bespoke or machine-learned metrics, target relatively common conditions, and do not preserve an established clinical scoring system. None addresses the ICARS template-tracing paradigm or rare ataxic disease. Consequently, there remains no digital platform designed for the ICARS template-tracing paradigm, limiting standardized quantitative assessment in pediatric rare disease natural history studies, multicenter clinical trials, and remote follow-up, where portable, low-burden digital biomarkers are particularly needed because of small, geographically dispersed patient populations.

Here we present and release ArchSpiral, an iPad application that digitizes the ICARS Archimedean spiral assessment while preserving its validated ordinal scoring. ArchSpiral generates automated ICARS-compatible scores and exportable quantitative tracing metrics. By combining ICARS-compatible scoring with digital biomarkers, ArchSpiral provides a standardized outcome assessment for longitudinal pediatric rare disease research, multicenter clinical trials, and telehealth-based follow-up while maintaining compatibility with the established ICARS scale. We evaluated ArchSpiral in 12 participants with CDG undergoing longitudinal natural history assessment.

### Objective

This study aimed to develop and implement an iPad-based application that digitizes the ICARS Archimedean spiral assessment in pediatric rare disease cohort while preserving clinical scoring and providing quantitative measures of spiral tracing performance.

## Materials and methods

### Application

#### Software Architecture and Spiral Acquisition

The ArchSpiral application was developed for iOS 26 using Swift, with SwiftUI providing the graphical user interface and PencilKit enabling high-resolution Apple Pencil input. The application was evaluated on a 9th-generation iPad using either finger touch or a stylus. The application renders a dynamically scaled three-turn Archimedean spiral template defined by the equation r=a+bθ, where the scaling parameter is calculated from the available canvas dimensions to maintain a geometrically consistent reference spiral across devices.

To standardize participant instruction, the application includes an animated demonstration overlay illustrating the expected tracing path before data acquisition, with the option to replay the demonstration before assessment. Spiral tracings, performed using either Apple Pencil or finger touch input, are captured as native PKDrawing objects, from which individual stroke points are extracted together with their spatial coordinates and timestamps.

#### Quantitative Spiral Analysis

Each sampled point is transformed from Cartesian to polar coordinates relative to the spiral center, and angular coordinates are unwrapped and normalized to generate a continuous trajectory. Tracing accuracy is quantified by uniformly resampling both the participant’s tracing and the reference Archimedean spiral and calculating the proportion of sampled points whose minimum Euclidean distance between the participant tracing and the reference Archimedean spiral falls within a predefined tolerance threshold, with distances evaluated in both trace-to-template and template-to-trace directions. Overall geometric accuracy is summarized using the root mean squared error (RMSE) derived from these bidirectional nearest-distance measurements. This bidirectional geometric comparison measures the participant’s adherence to the mathematically defined Archimedean spiral while also accounting for coverage of the reference trajectory.

Additional performance metrics include drawing duration, average drawing speed calculated as total path length divided by elapsed time, finger/pen lift count determined from the number of discrete PencilKit strokes, and pause count based on temporal gaps between consecutive sampled points. Drawing smoothness is quantified using a steadiness metric calculated from the residual radial error following subtraction of a moving-average trend, isolating short-scale trajectory irregularity independent of the underlying spiral geometry. Higher steadiness scores indicate smoother tracing.

#### Automated ICARS-Compatible Spiral Morphology Scoring

The application additionally implements an automated ICARS-compatible spiral morphology score. For scoring, the traced spiral is downsampled by removing closely spaced sampled points while preserving the overall tracing trajectory, and the resulting line segments are analyzed using computational geometry algorithms to identify self-touching, overlap, and true self-crossings using line-segment intersection testing, minimum segment-to-segment distance calculations, angular separation between spiral loops, and crossing-angle thresholds while excluding contacts between adjacent or nearby portions of the same spiral loop to minimize false-positive detections.

#### Data Export and Reporting

Upon completion, ArchSpiral automatically generates quantitative outputs including tracing accuracy, RMSE, tolerance threshold, drawing duration, average drawing speed, finger/pen lift count, pause count, steadiness score, and automated ICARS spiral score. Results can be exported through the iOS sharing interface as a structured text report containing all quantitative metrics, a rendered PNG image, and the native PKDrawing file for archival, sharing, or subsequent reanalysis. Assessment data are stored locally on the iPad in a de-identified format and are not automatically uploaded to cloud storage (Figure 1).

**Figure 1.**
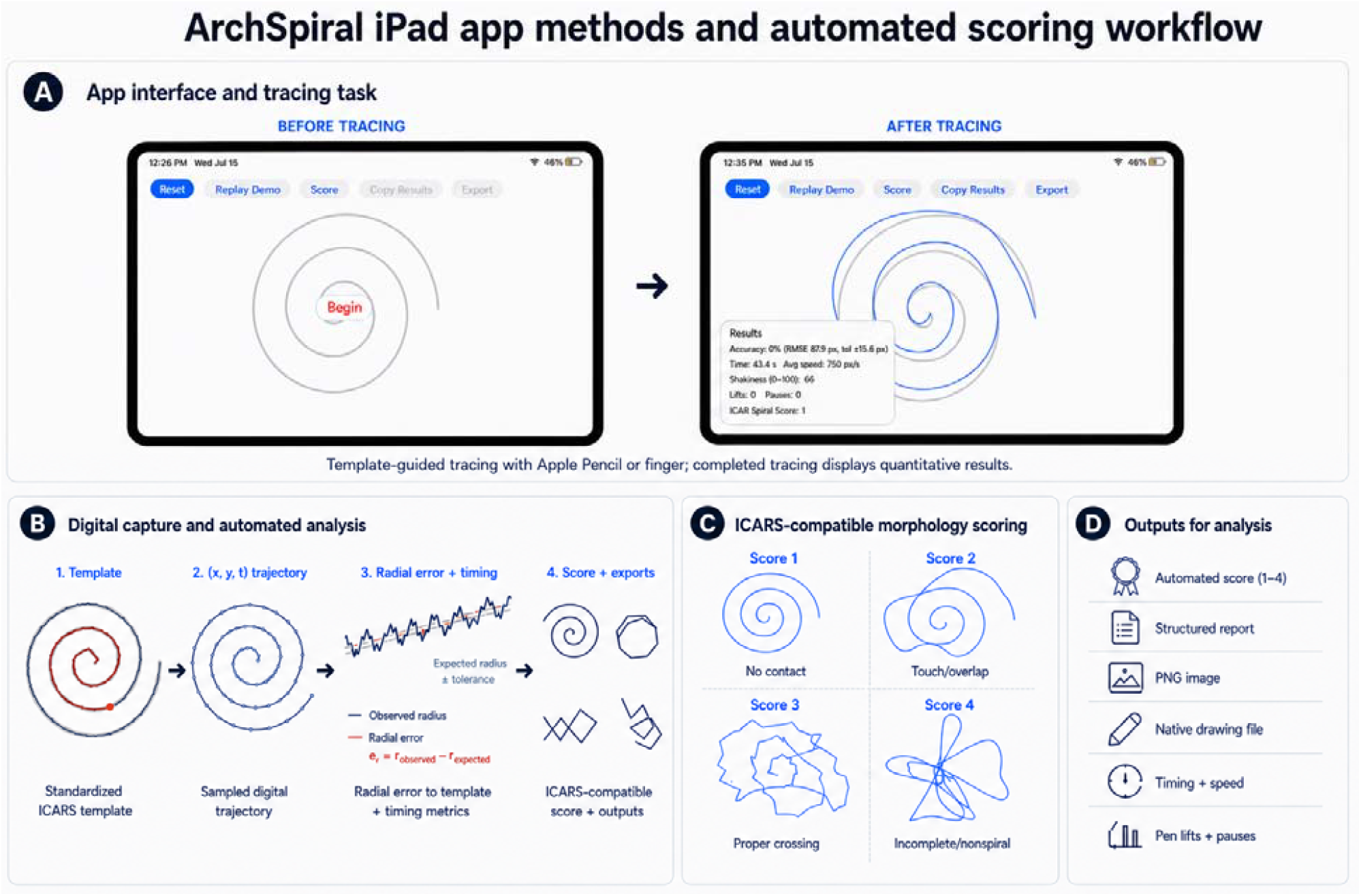
ArchSpiral iPad application workflow and automated analysis pipeline. **(A)** ArchSpiral application interface before and after the digital Archimedean spiral assessment. **(B)** Automated digital analysis workflow from standardized spiral tracing and trajectory capture to bidirectional geometric distance analysis, quantitative metric generation, and automated ICARS-compatible spiral scoring. **(C)** Representative examples of the four automated ICARS-compatible spiral morphology scoring categories. **(D)** Quantitative outputs generated by the application, including automated ICARS-compatible spiral score, structured results report, PNG image, native drawing file, drawing time and speed, and pen/finger lift and pause metrics.

### Cohort

We evaluated 12 individuals with clinically and molecularly confirmed CDG enrolled in the CDG Natural History Protocol (IRB: 19-016991_RS03). Participants completed the paper-based ICARS Archimedean spiral tracing task, followed by the corresponding ArchSpiral assessment on an iPad using either stylus or finger input.

Collected metrics included total ICARS score, kinetic function subscore, paper-based and digital ICARS spiral scores, tracing accuracy (%), RMSE (pixels), tolerance threshold (pixels), drawing duration (seconds), average drawing speed (pixels/second), steadiness score (0–100; higher scores indicate smoother tracing), pen lift count (n), and pause count (n).

### Statistical Analysis

Paper-based and digital ICARS spiral scores, ordinal (1–4), were compared using Spearman rank correlation. Agreement was summarized by exact agreement, agreement within one ordinal category, and linear weighted Cohen’s κ. Seven continuous digital drawing metrics (accuracy, RMSE, drawing duration, average drawing speed, steadiness, pen lifts, and pauses) were correlated with the paper-based spiral score and, as an internal consistency assessment, with the algorithm-derived digital spiral score, applying Benjamini–Hochberg false-discovery-rate correction across the seven metrics. Statistical analyses were performed in Python using SciPy and statsmodels.

## Results

### Cohort and Clinical Evaluation

Participants who completed the ArchSpiral assessment ranged in age from 5 to 39 years, with a mean age of 21.1 years. Eight participants (66.7%) were pediatric, defined as younger than 21 years. The cohort represented five CDG subtypes: PMM2-CDG (n = 5), DHDDS-CDG (n = 2), NUS1-CDG (n = 2), ALG2-CDG (n = 2), and DPGAT1-CDG (n = 1). Total ICARS scores ranged from 11 to 89, kinetic function subscores from 7 to 48, and paper-based ICARS spiral scores from 0 to 4.

### Quantitative spiral analysis

ArchSpiral generated quantitative measurements for all participants. Tracing accuracy ranged from 0% to 46%, RMSE from 24.7 to 240.7 pixels, drawing duration from 7.46 to 77.82 seconds, average drawing speed from 97 to 875 pixels/second, steadiness from 0 to 99, pen lift count from 0 to 19, and pause count from 0 to 4. Digital ICARS spiral scores spanned the full scoring range (1–4), demonstrating classification across a spectrum of motor impairment (Figure 1).

### Comparison with conventional ICARS scoring

Paper-based and digital ICARS spiral scores agreed exactly in 11 of 12 participants (91.7%) and within one ordinal category in all participants (linear weighted Cohen’s κ = 0.91), differing by a single category in one participant (Figure 2). Among the quantitative digital metrics, steadiness showed the strongest association with the paper-based spiral score (Spearman’s ρ = −0.82) and was the only metric to remain significant after false-discovery-rate correction (adjusted *P* = 0.008). RMSE (ρ = 0.61) and pen lifts (ρ = 0.59) trended in the expected direction but did not remain significant after correction. Correlations with the automated digital spiral score were similar in magnitude and were interpreted as an internal consistency assessment (Figure 2B–F; Table 2).

**Table 1.** Participant-level paper and digital Archimedean spiral assessment results. Paper-based International Cooperative Ataxia Rating Scale (ICARS) spiral assessments were completed before digital ArchSpiral assessment. Conventional paper spiral scores were compared with automated digital spiral scores and quantitative metrics generated by ArchSpiral, including tracing accuracy (%), root mean squared error (RMSE, px), tolerance threshold (px), drawing duration (s), average drawing speed (px/s), steadiness score, pen lift count, and pause count.

| Participant | Diagnosis | Paper Spiral Score | Accuracy (%) | RMSE (px) | Tolerance (px) | Time (s) | Avg Speed (px/s) | Steadiness | Lifts | Pauses | Digital ICARS Spiral Score |
| --- | --- | --- | --- | --- | --- | --- | --- | --- | --- | --- | --- |
| 1 | DHDDS | 1 | 21 | 34.1 | ±16.2 | 30.92 | 97 | 95 | 1 | 1 | 1 |
| 2 | DHDDS | 1 | 0 | 103.7 | ±16.2 | 14.85 | 264 | 76 | 0 | 0 | 1 |
| 3 | NUS1 | 1 | 44 | 24.7 | ±16.2 | 15.47 | 261 | 88 | 0 | 0 | 1 |
| 4 | NUS1 | 1 | 0 | 75.7 | ±16.2 | 11.91 | 384 | 74 | 1 | 0 | 1 |
| 5 | PMM2 | 3 | 0 | 240.7 | ±16.2 | 77.82 | 99 | 55 | 4 | 4 | 3 |
| 6 | PMM2 | 1 | 0 | 90.8 | ±16.2 | 36.29 | 100 | 95 | 0 | 0 | 1 |
| 7 | ALG2 | 1 | 39 | 36.3 | ±16.2 | 19.89 | 186 | 88 | 1 | 0 | 1 |
| 8 | ALG2 | 2 | 6 | 103.9 | ±16.2 | 9.17 | 414 | 28 | 2 | 0 | 2 |
| **9** | DPGAT1 | 1 | 27 | 39.6 | ±16.2 | 12.28 | 308 | 91 | 0 | 0 | 1 |
| **10** | PMM2 | 3 | 35 | 36.9 | ±16.2 | 30.47 | 142 | 71 | 0 | 0 | 2 |
| **11** | PMM2 | 4 | 0 | 196.5 | ±16.2 | 7.46 | 875 | 0 | 19 | 0 | 4 |
| **12** | PMM2 | 1 | 46 | 26.9 | ±16.2 | 22.18 | 171 | 99 | 0 | 0 | 1 |

**Table 2.**
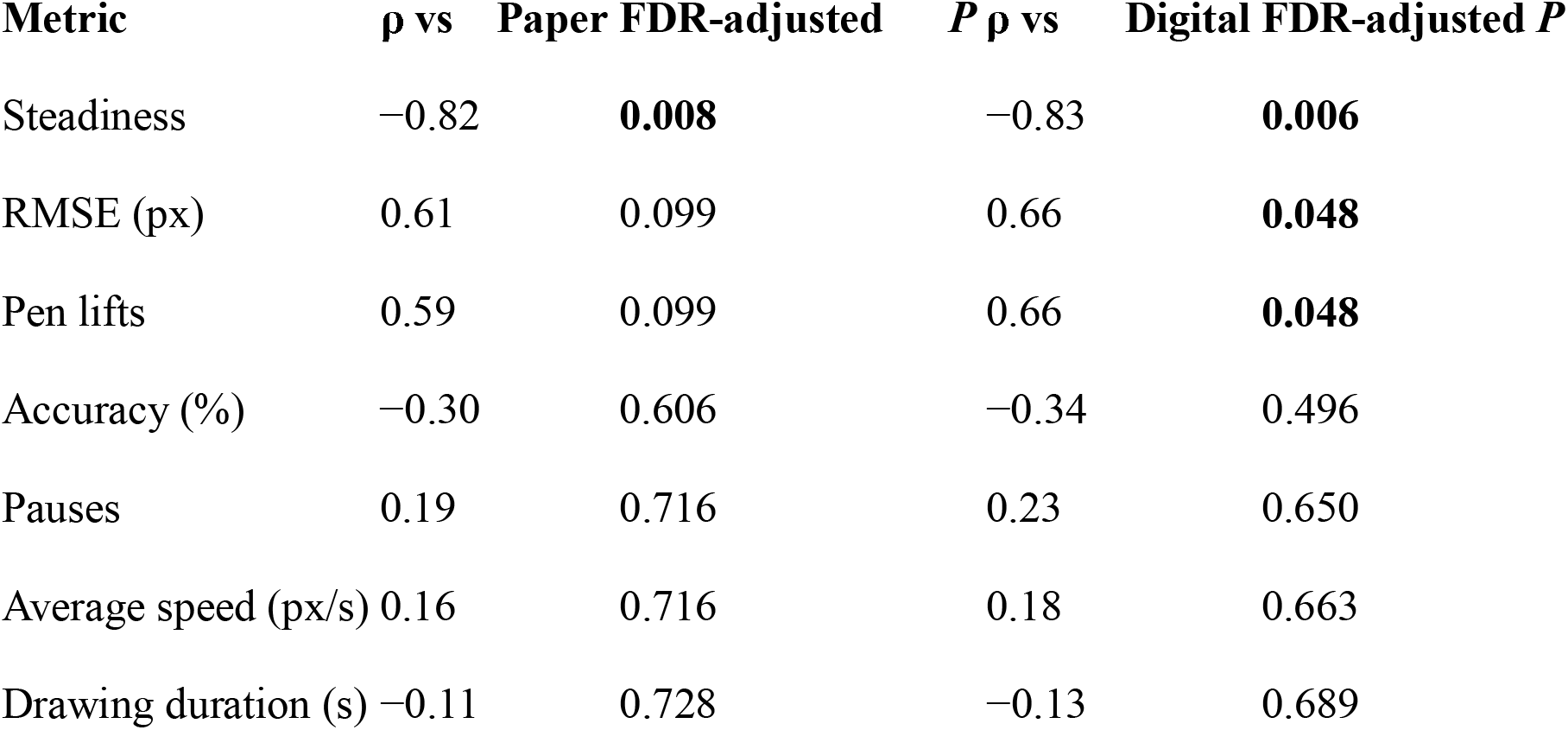
Correlation of quantitative digital spiral metrics with paper-based and automated digital International Cooperative Ataxia Rating Scale (ICARS) spiral scores. Spearman rank correlation coefficients (ρ) and false-discovery-rate (FDR)-adjusted *P* values are shown for each quantitative digital metric relative to the paper-based and automated digital ICARS spiral scores. Benjamini–Hochberg false-discovery-rate correction was applied across the seven metrics for each comparison. **\* Bold** = survives FDR < 0.05. RMSE and pen lifts survive against the digital score but not against the paper score. Because the digital score is algorithm-derived and its feature overlap with these metrics is unknown, the apparently stronger performance may be partly built in rather than independent; these correlations should be interpreted as an internal consistency assessment, not confirmation.

| **Metric** | **$\rho$ vs Paper** | **FDR-adjusted $P$** | **$\rho$ vs Digital** | **FDR-adjusted $P$** |
| --- | --- | --- | --- | --- |
| Steadiness | −0.82 | **0.008** | −0.83 | **0.006** |
| RMSE (px) | 0.61 | 0.099 | 0.66 | **0.048** |
| Pen lifts | 0.59 | 0.099 | 0.66 | **0.048** |
| Accuracy (%) | −0.30 | 0.606 | −0.34 | 0.496 |
| Pauses | 0.19 | 0.716 | 0.23 | 0.650 |
| Average speed (px/s) | 0.16 | 0.716 | 0.18 | 0.663 |
| Drawing duration (s) | −0.11 | 0.728 | −0.13 | 0.689 |

**Figure 2.**
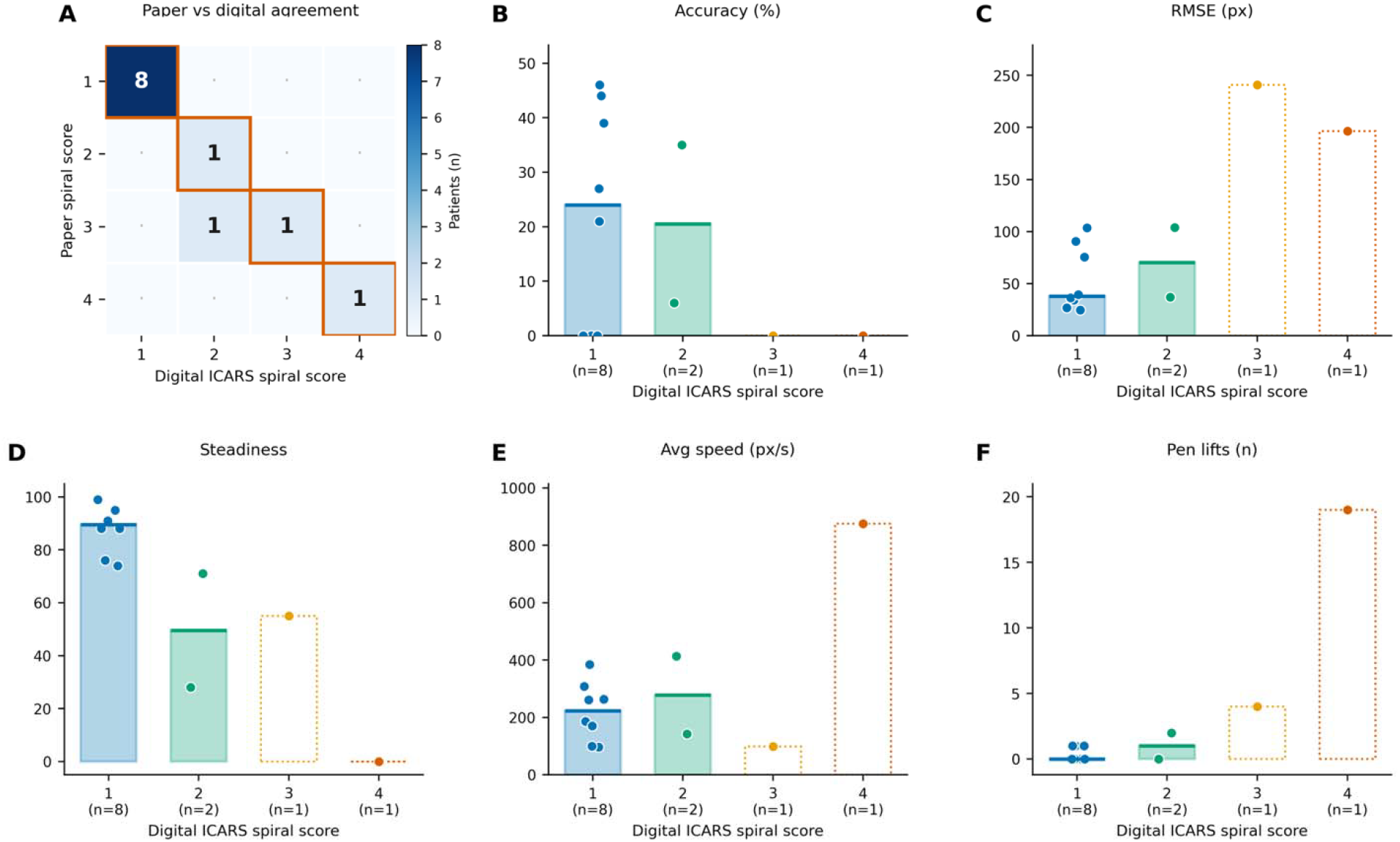
Comparison of conventional paper-based International Cooperative Ataxia Rating Scale (ICARS) spiral scoring and quantitative digital ArchSpiral metrics. (A) Agreement between paper-based and automated digital ICARS spiral scores. (B–F) Quantitative digital metrics stratified by automated digital ICARS spiral score, including tracing accuracy (%), root mean squared error (RMSE, px), steadiness score, average drawing speed (px/s), and pen lift count (n). Individual participant values are shown with group means. Higher steadiness scores indicate smoother tracing.

## Discussion

This study demonstrates the successful implementation of an iPad-based Archimedean spiral application in a CDG natural history cohort while preserving the established ICARS spiral assessment used for longitudinal disease monitoring. ArchSpiral complements conventional ICARS scoring with continuous quantitative biomarkers. Because the ICARS spiral assessment is a 0–4 scale, meaningful differences in tracing quality and motor performance may exist among participants assigned the same score or within the same participant over time. These continuous measures may detect subtle changes in motor performance that would not otherwise result in a change in the ordinal ICARS spiral score.

Agreement between paper-based and digital ICARS spiral scores demonstrates that the digital implementation faithfully reproduces the conventional clinical assessment. Exact agreement was observed in 11 of 12 assessments (91.7%), with all remaining assessments differing by only a single ordinal category. In addition, steadiness showed the strongest association with the paper-based ICARS spiral score and was the only quantitative metric that remained significant after false-discovery-rate correction. Together, these findings support the incorporation of ArchSpiral into existing ICARS-based assessments while extending conventional scoring through automated ICARS-compatible scoring and quantitative biomarkers.

Unlike most previously reported digital spiral assessment platforms, which have focused on freehand spiral drawing in common neurologic disorders, ArchSpiral was designed to digitize the standard ICARS template-tracing assessment while preserving ICARS scoring. This approach enables quantitative biomarker collection without requiring modification of existing clinical protocols or outcome measures used in rare disease natural history studies.

The feasibility of implementing ArchSpiral as an iOS-based application using either an Apple Pencil or finger touch may facilitate standardized remote assessment while capturing biomarkers of performance. This approach may be particularly valuable for pediatric rare disease populations, who often travel long distances to access specialized medical genetics care. A recent workforce study reported a mean travel distance of 76.6 miles for patients seeking medical genetics services, with some traveling as far as 386 miles.[16] By enabling standardized quantitative motor assessment on a widely available tablet platform while preserving ICARS scoring, ArchSpiral has the potential to expand the clinical information obtained during telehealth visits, reduce travel burden, and support longitudinal follow-up.

### Limitations

As a pilot implementation study, this study has several limitations. First, the cohort consisted exclusively of individuals with CDG, precluding comparisons with other populations. Future studies including healthy controls and individuals with other movement disorders are needed to establish normative ranges and determine the relationship of the quantitative biomarkers to disease severity, progression, and therapeutic response. Additionally, the digital assessment was completed using either finger touch or a stylus. Future studies should evaluate biomarker reproducibility across input modalities and determine whether one approach provides greater sensitivity for longitudinal assessment. To facilitate future studies, the application has since been updated to include an option to record whether finger touch or Apple Pencil input was used for each assessment.

## Conclusion

Our study demonstrates the successful implementation of an iPad-based application that digitizes the ICARS Archimedean spiral assessment while preserving its established clinical scoring system and generating quantitative measures of performance. By combining compatibility with an established clinical instrument and automated digital biomarker extraction, ArchSpiral provides a scalable, accessible platform for longitudinal monitoring and remote assessment, extending the validated ICARS assessment with objective measures that may improve longitudinal phenotyping and outcome assessment in pediatric rare disease research and clinical care.

## Data Availability

The Children's Hospital of Philadelphia IRB (IRB: 19-016991_RS03) gave ethical approval for this work.

## Author contributions

Daniel Schecter (Conceptualization, Software, Investigation, Methodology, Writing—original draft), Rory Tinker (Methodology, Writing— review & editing), Matteo Danieletto (Methodology, Writing—review & editing, Software), Georgia MacDonald (Methodology, Writing— review & editing), Tamas Kozicz (Investigation, Methodology, Writing—review & editing), Eva Morava (Investigation, Methodology, Supervision, Writing—review & editing), and Benjamin Glicksberg (Investigation, Methodology, Supervision, Writing— review & editing).

## Funding

This work was supported in part through the Minerva computational and data resources and staff expertise provided by Scientific Computing and Data at the Icahn School of Medicine at Mount Sinai and supported by the Clinical and Translational Science Awards (CTSA) grant UL1TR004419 from the National Center for Advancing Translational Sciences. This work was also supported by the Hasso Plattner Foundation. The CDG Natural History study was supported by the Frontiers in Congenital Disorders of Glycosylation grant (U54NS115198) from the National Institute of Neurological Disorders and Stroke (NINDS), the National Center for Advancing Translational Sciences (NCATS), the Eunice Kennedy Shriver National Institute of Child Health and Human Development (NICHD), and the Rare Diseases Clinical Research Network (RDCRN) of the National Institutes of Health. R.J.T. was supported by a T32GM082773. We thank CDG CARE for its continued support and all the patients and families who participated in this study.

## Conflicts of interest

The authors have no relevant competing interest to share.

## Notes

### Competing Interest Statement

The authors have declared no competing interest.

### Author Declarations

Ethics committee/IRB (CDG Natural History Protocol; IRB: 19-016991_RS03) of Children's Hospital of Philadelphia gave ethical approval for this work

